# The Burnout Syndrome among Dietitians in Arar, Saudi Arabia

**DOI:** 10.1101/2024.01.30.24301842

**Authors:** Nasser Alqahtani, Bushra Albadareen, Amna Mohammad-Mahmood

## Abstract

This study identified and described the status of burnout experienced by Saudi dietitians in various settings. A questionnaire was sent to 160 members who were invited to participate via informal networks. The Copenhagen Burnout Inventory (CBI) scale was used to measure burnout status. The CBI comprises three subscales: personal burnout **PB** (six items), work burnout **WB** (seven items), and client-related burnout **CB** (six items). Among the 160 participants in this study, 94 participants (58.75%) experienced burnout. most of the participants had moderate PB (n = 49, 30.6%), low WB (n = 58, 36.3%), and moderate CB (n = 73, 45.6%). These results suggest that the Saudi dietitians as a group perceive themselves to be moderately personal and client-related burnout, and to have a low level of work burnout. Further analysis also suggests that Saudi dietitians in nontraditional settings experience more burnout than those in healthcare clinical settings.

## I. INTRODUCTION

The concept of burnout, as “a syndrome of emotional exhaustion, depersonalization, and reduced personal accomplishment that can occur as the result of interaction between different personal and professional factors (1), can affect health, giving rise to both physical and psychosomatic problems as well as depression, anxiety, low self-esteem, guilt feelings, and low tolerance of frustration (2-4). In the field of health, healthcare personnel have frequently been the target of study to measure the repercussions of their work on their own health. A number of studies have found that ongoing exposure to occupational stress can lead to burnout syndrome and that this can affect physical, psychological, and social health (5-7). Dietitians are integral members of the healthcare team. However, many dietitians express that their acceptance as full healthcare team members is variable. Having their work undermined or ignored has been a source of frustration and is believed to be a major cause of stress and burnout among dietitians (8). For that, we investigated the prevalence of burnout among Saudi dietitians and established an association with Sociodemographic, academic, and practical-related factors.

## II. MATERIALS AND METHODS

### Design

A cross-sectional survey design was conducted from February to April 2021 in Arar, Saudi Arabia.

### Sample and recruitment

Approximately 160 Saudi dietitians were invited to participate via informal networks. The online survey included an invitation outlining the aims of the study, contact details of researchers, and a live link to the host survey platform. Measures The full survey included demographic questions and the Copenhagen Burnout Inventory (CBI) scales. The CBI comprises three subscales: personal burnout **PB** (six items), work burnout **WB** (seven items), and client-related burnout **CB** (six items). Twelve items have responses of frequency along a five-point scale ranging from 100 (always), 75 (often), 50 (sometimes), 25 (seldom), and 0 (never/almost never). Seven items use response categories according to intensity ranging from a very low degree to a very high degree. Scores of 50 to 74 are considered ‘moderate’, 75–99 are high, and a score of 100 is considered severe burnout. All items are straightforward, positively skewed, relate to the relevant subscale, and have high internal reliability (9).

### Data availability

The data supporting this study’s findings are available from the corresponding author, upon reasonable request.

#### Sample Size Calculation

According to the Raosoft online sample size calculator, to achieve a confidence level of 95% with an error margin of 5%, a minimum sample size of 160 was required to accomplish the study’s aims.

### Statistical analyses

Analysis was done using SPSS version 16.5 (Chicago, IL, USA). Data regarding the socio-demographic characteristics, and academic, and practical-related factors of participants were obtained. ANOVA and independent t-test were used to compare the means of each subscale of CBI to the socio-demographic, academic, and practical-related factors. Chi-square was used to compare the classification of CBI subscale scores and the presence of burnout with socio-demographic, academic, and practical-related factors. Results were considered statistically significant at a *p*-value < 0.05.

## III. RESULTS

A total of 160 participants completed the questionnaire. The detailed characteristics of study participants are presented in **Table 1**. The participants were predominantly 18-29 years old (76.3 %), female (81.9 %), bachelor’s degree (81.3%), with less than 10.000 SR per month (60%). Most of the participants practiced in the clinical field (81.9%), with a 1-5 year duration (55%). **Table 2** presents the comparison between the mean of each subscale score and all the identified factors. There was a statistically significant difference upon a comparison of the mean of each subscale score (PB, WB, and CB) with age, salary, practical area, and years (p = 0.00), with no significant difference upon a comparison of mean each subscale scores (PB, WB, and CB) with gender and academic level. The lower burnout score for each subscale was found in 18 -29-year-old participants with a salary < 10.000 SR who are working in the clinical field and experience of less than 1 year. **Table 3** presents the overall subscale scores of the residents in this study. In this study, most of the participants had moderate PB (n = 49, 30.6%), low WB (n = 58, 36.3%), and moderate CB (n = 73, 45.6%). **Table 4** contains the comparison between all the subscale scores and the evaluated factors. Upon comparison between all subscale classifications and socio-demographic, academic, and work-related factors. Most of the participants were aged (18 - 29) years, Salary < 10,000 SR, clinical field, and (1-5) practical years had a slightly lower to moderate tendency to have PB, WB, and CB. **Table 5** compares the presence of burnout and the related factors. Among the 160 participants in this study, 94 participants (58.75%) experienced burnout. There were statistically significant differences in age, salary, practical area, and years (p = 0.00), upon comparison with the absence of burnout. According to age (18 - 29) years 57 (46.7%), Salary <10.000 SR 39 (40.6%), clinical field 68 (51.9%) and (1-5) practical years 10 (24.4%) were found to experience more burnout.

**Table 1.**
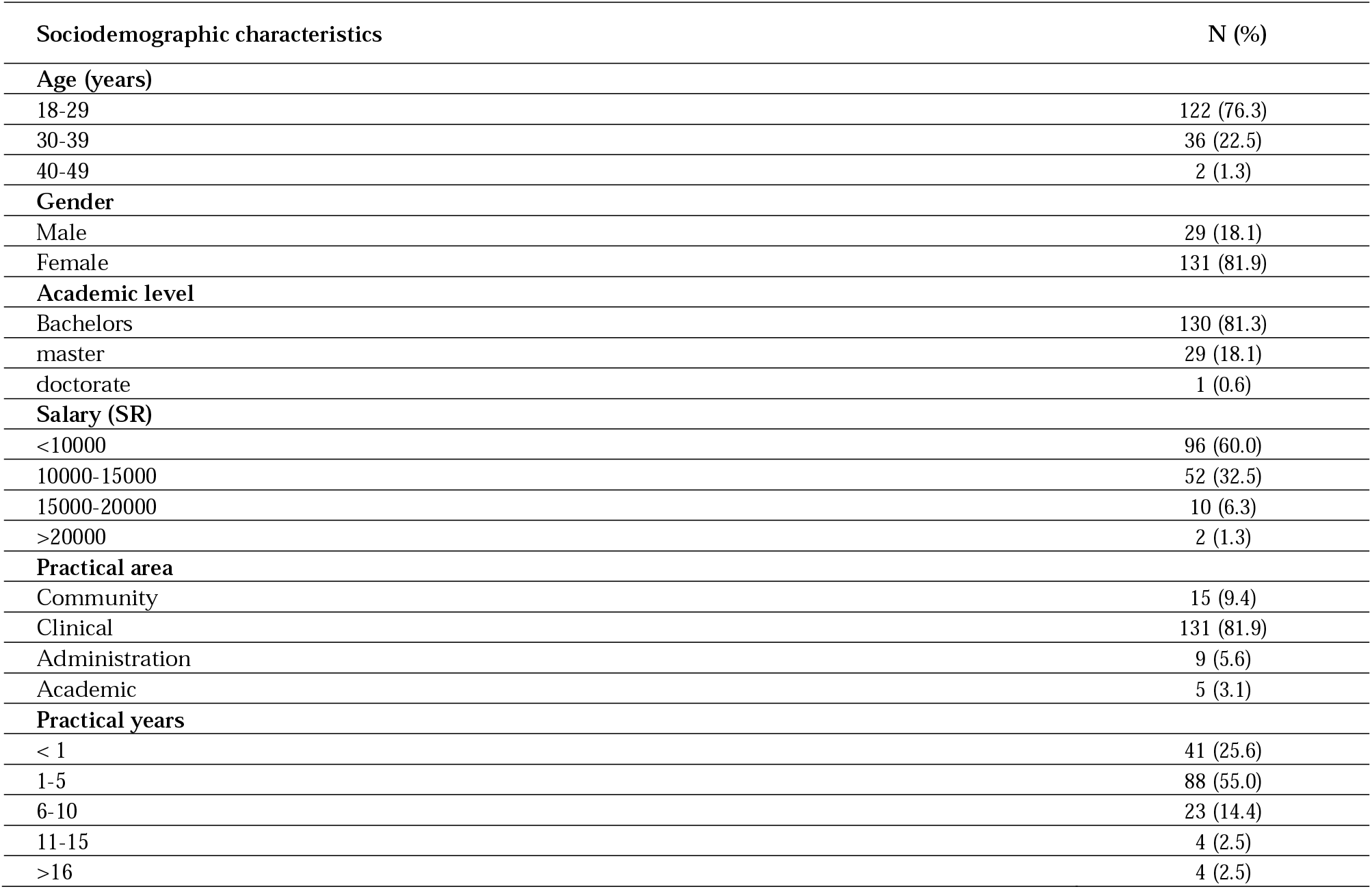
Socio-demographic characteristics, academic, and practical-related factors of study participants.

**Table 2.**
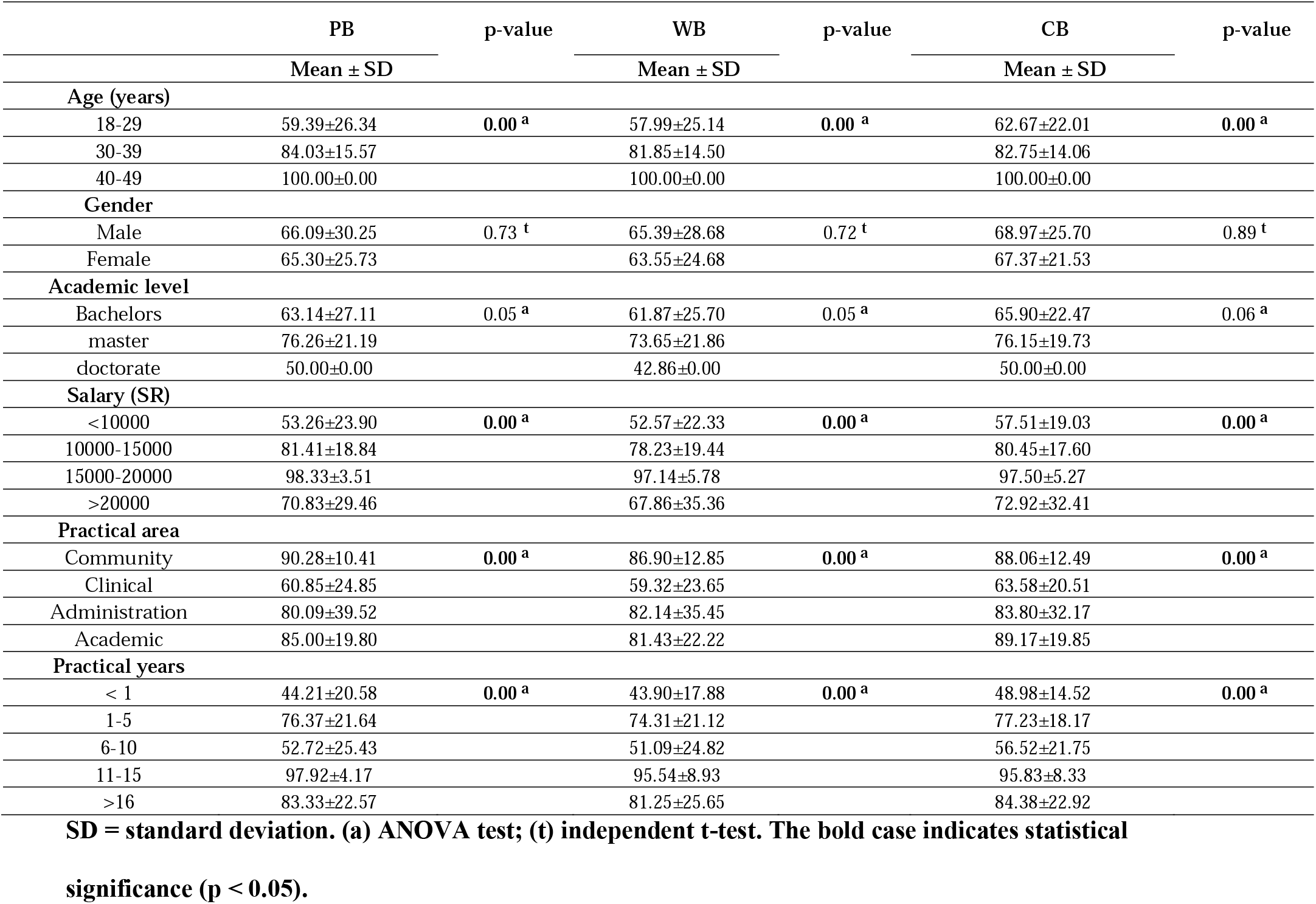
Comparison between the mean of each subscale scores of CBI and socio-demographic, academic, and practical-related factors.

|  | **PB** | **p-value** | **WB** | **p-value** | **CB** | **p-value** |
| --- | --- | --- | --- | --- | --- | --- |
|  | **Mean ± SD** |  | **Mean ± SD** |  | **Mean ± SD** |  |
| **Age (years)** |  |  |  |  |  |  |
| 18-29 | 59.39±26.34 | **0.00<sup>a</sup>** | 57.99±25.14 | **0.00<sup>a</sup>** | 62.67±22.01 | **0.00<sup>a</sup>** |
| 30-39 | 84.03±15.57 |  | 81.85±14.50 |  | 82.75±14.06 |  |
| 40-49 | 100.00±0.00 |  | 100.00±0.00 |  | 100.00±0.00 |  |
| **Gender** |  |  |  |  |  |  |
| Male | 66.09±30.25 | 0.73<sup>t</sup> | 65.39±28.68 | 0.72<sup>t</sup> | 68.97±25.70 | 0.89<sup>t</sup> |
| Female | 65.30±25.73 |  | 63.55±24.68 |  | 67.37±21.53 |  |
| **Academic level** |  |  |  |  |  |  |
| Bachelors | 63.14±27.11 | 0.05<sup>a</sup> | 61.87±25.70 | 0.05<sup>a</sup> | 65.90±22.47 | 0.06<sup>a</sup> |
| master | 76.26±21.19 |  | 73.65±21.86 |  | 76.15±19.73 |  |
| doctorate | 50.00±0.00 |  | 42.86±0.00 |  | 50.00±0.00 |  |
| **Salary (SR)** |  |  |  |  |  |  |
| <10000 | 53.26±23.90 | **0.00<sup>a</sup>** | 52.57±22.33 | **0.00<sup>a</sup>** | 57.51±19.03 | **0.00<sup>a</sup>** |
| 10000-15000 | 81.41±18.84 |  | 78.23±19.44 |  | 80.45±17.60 |  |
| 15000-20000 | 98.33±3.51 |  | 97.14±5.78 |  | 97.50±5.27 |  |
| >20000 | 70.83±29.46 |  | 67.86±35.36 |  | 72.92±32.41 |  |
| **Practical area** |  |  |  |  |  |  |
| Community | 90.28±10.41 | **0.00<sup>a</sup>** | 86.90±12.85 | **0.00<sup>a</sup>** | 88.06±12.49 | **0.00<sup>a</sup>** |
| Clinical | 60.85±24.85 |  | 59.32±23.65 |  | 63.58±20.51 |  |
| Administration | 80.09±39.52 |  | 82.14±35.45 |  | 83.80±32.17 |  |
| Academic | 85.00±19.80 |  | 81.43±22.22 |  | 89.17±19.85 |  |
| **Practical years** |  |  |  |  |  |  |
| < 1 | 44.21±20.58 | **0.00<sup>a</sup>** | 43.90±17.88 | **0.00<sup>a</sup>** | 48.98±14.52 | **0.00<sup>a</sup>** |
| 1-5 | 76.37±21.64 |  | 74.31±21.12 |  | 77.23±18.17 |  |
| 6-10 | 52.72±25.43 |  | 51.09±24.82 |  | 56.52±21.75 |  |
| 11-15 | 97.92±4.17 |  | 95.54±8.93 |  | 95.83±8.33 |  |
| >16 | 83.33±22.57 |  | 81.25±25.65 |  | 84.38±22.92 |  |

**SD = standard deviation. (a) ANOVA test; (t) independent t-test. The bold case indicates statistical**

**Table 3.**
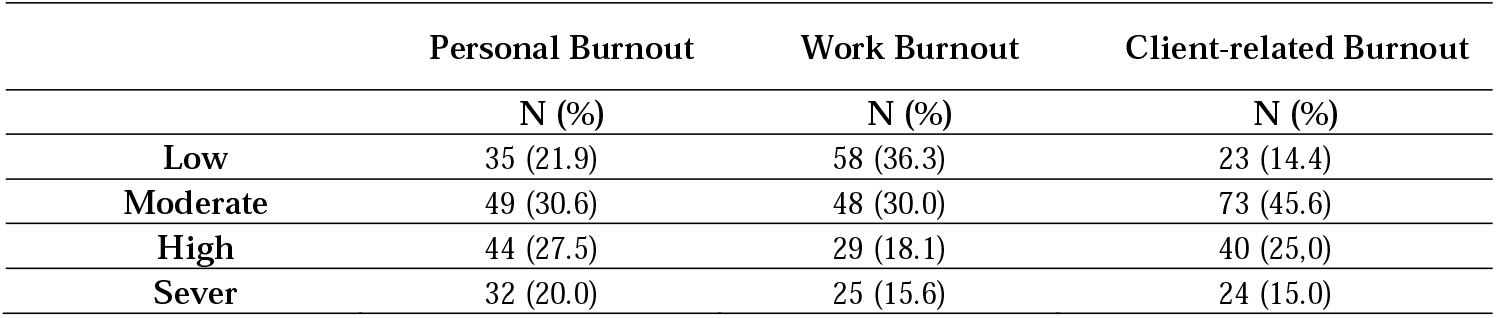
Overall personal, work, and client-related burnout subscale scores of participants in the study.

|  | Personal Burnout | Work Burnout | Client-related Burnout |
| --- | --- | --- | --- |
|  | N (%) | N (%) | N (%) |
| **Low** | 35 (21.9) | 58 (36.3) | 23 (14.4) |
| **Moderate** | 49 (30.6) | 48 (30.0) | 73 (45.6) |
| **High** | 44 (27.5) | 29 (18.1) | 40 (25.0) |
| **Sever** | 32 (20.0) | 25 (15.6) | 24 (15.0) |

**Table 4.**
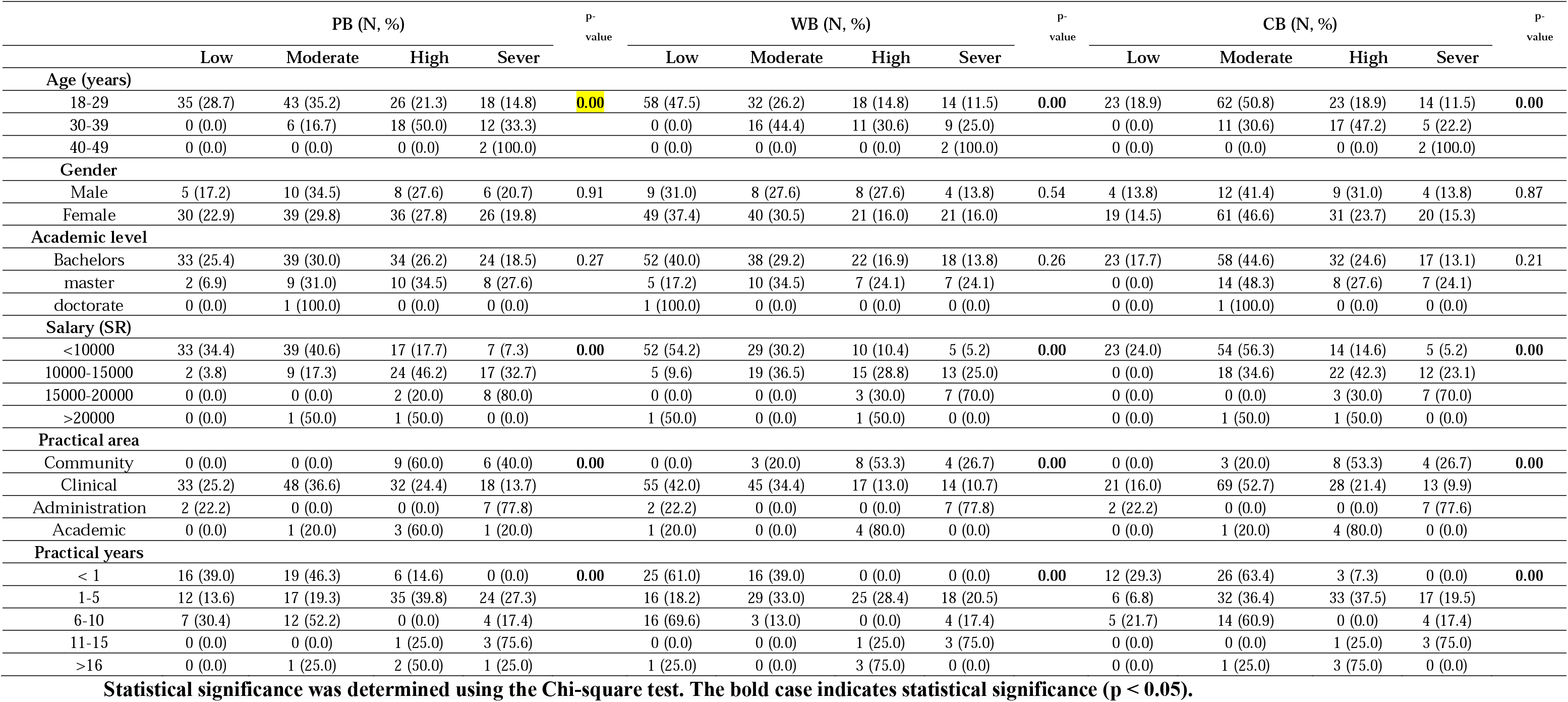
Comparison between CBI subscale scores and socio-demographic, academic, and practical-related factors.

**Table 5.** Comparison between the absence and presence of burnout and socio-demographic, academic, and practical-related factors.

| BURNOUT (N, %) |  |  |  |
| --- | --- | --- | --- |
|  | Absent | Present | p-value |
| Age (years) |  |  |  |
| 18-29 | 65 (53.3) | 57 (46.7) | 0.00 |
| 30-39 | 1 (2.8) | 35 (97.2) |  |
| 40-49 | 0 (0.0) | 2 (100) |  |
| Gender |  |  |  |
| Male | 10 (34.5) | 19 (65.5) | 0.27 |
| Female | 56 (42.7) | 75 (57.3) |  |
| Academic level |  |  |  |
| Bachelors | 57 (43.8) | 73 (56.2) | 0.13 |
| master | 8 (27.6) | 21 (72.4) |  |
| doctorate | 1(100.0) | 0 (0.0) |  |
| Salary (SR) |  |  |  |
| <10000 | 57 (59.4) | 39 (40.6) | 0.00 |
| 10000-15000 | 8 (15.4) | 44 (84.6) |  |
| 15000-20000 | 0 (0.0) | 10 (100.0) |  |
| >20000 | 1 (50.0) | 1 (50.0) |  |
| Practical area |  |  |  |
| Community | 0 (0.0) | 15 (100.0) | 0.00 |
| Clinical | 63 (48.1) | 68 (51.9) |  |
| Administration | 2 (22.2) | 7 (77.8) |  |
| Academic | 1 (20.0) | 4 (80.0) |  |
| Practical years |  |  |  |
| < 1 | 31 (75.6) | 10 (24.4) | 0.00 |
| 1-5 | 16 (18.2) | 72 (81.8) |  |
| 6-10 | 18 (78.3) | 5 (21.7) |  |
| 11-15 | 0 (0.0) | 4 (100.0) |  |
| >16 | 1 (25.0) | 3 (75.0) |  |

Statistical significance was determined using the Chi-square test. The bold case indicates statistical significance ( $p < 0.05$ ).

## IV. DISCUSSION

This cross-sectional survey of Saudi dietitians revealed moderate personal, work, and client-related burnout. Furthermore, Among the 160 participants in this study, 94 participants (58.75%) experienced burnout. To the best of our knowledge, the present study is the first to assess the prevalence of burnout and burnout-associated factors among Saudi dietitians. Burnout and its consequences cost employees and organizations. Burnout has negative impacts on the well-being of healthcare professionals and increases the risk of job withdrawal, absenteeism, and suicidal ideation (10-13). The results showed a slightly lower to moderate tendency to have PB, WB, and CB in participants aged (18 - 29) years, with a Salary <10.000 SR, worked in the healthcare clinical field, and had (1-5) practical years.

Therefore, it can be concluded that dietitians who are older than 30 years old are more likely to experience higher levels of burnout than younger than 30. The other significant associations were that low-salary workers had lower levels of burnout than high-salary workers. On the other hand, the more practical years dietitians have, the more likely they will suffer burnout. However, Buick and Thomas (14) identified the profile who are most likely to experience burnout as relatively young, single or divorced, female, and relatively new to managerial responsibilities. It is true that demographic and organizational characteristics can be personal and situational factors that influence job burnout perceptions (15). Many healthcare sites employ dietitians in positions that do not require their skills or certification even though the title is for dietitians. The comments by the respondents suggested that superiors and other staff members do not appreciate or fully utilize the skills they offer. Such underutilization was reported as early as 1981 (16). Therefore, recognizing and utilizing dietitians to their fullest potential would assist in overcoming this problem. Work settings were found to be an important factor in increasing perceptions of personal accomplishment and reduced PB. Our results showed that dietitians who reported working in nontraditional settings had significantly lower personal accomplishment perceptions and higher PB than those working in healthcare clinical settings. These dietitians may be considered high achievers, recognizing their ability and moving into traditional areas.

## V. LIMITATION

There are several limitations to the present study, including the use of a non-random sampling technique, as these may not be representatives of the entire dietitian population. Furthermore, as a cross-sectional survey is an observational study design, no causation can be drawn from the results.

## VI. CONCLUSION

The main findings were the following. Age, salary, practical area, and years were the variables significantly associated with the levels of burnout experienced by Saudi dietitians. It also appeared that those dietitians who are older than 30 years old are more likely to experience higher levels of burnout than younger than 30. The other significant associations were that low-salary workers had lower levels of burnout than high-salary workers. On the other hand, the more practical year’s dietitians have, the more likely they will suffer burnout. Dietitians who reported working in nontraditional settings had significantly lower personal accomplishment perceptions and higher PB than those working in healthcare clinical settings.

## Acknowledgments

We would like to thank the participants who volunteered to be enrolled in this study.

